# Effectiveness of COVID-19 mRNA vaccine booster dose relative to primary series during a period of Omicron circulation

**DOI:** 10.1101/2022.04.15.22273915

**Authors:** Joshua G. Petrie, Jennifer P. King, David L. McClure, Melissa A. Rolfes, Jennifer K. Meece, Edward A. Belongia, Huong Q. McLean

**Affiliations:** Marshfield Clinic Research Institute, Marshfield, Wisconsin, USA; Influenza Division, Centers for Disease Control and Prevention, Atlanta, Georgia, USA

**Keywords:** SARS-CoV-2, COVID-19, Omicron, Vaccine Effectiveness, Booster

## Abstract

During a period of Omicron variant circulation, we estimated relative VE of COVID-19 mRNA booster vaccination versus primary two-dose series in an ongoing community cohort. Relative VE was 66% (95% CI: 46%, 79%) favoring the booster dose compared to primary series vaccination. Our results support current booster recommendations.

---

Clinical trials supporting emergency use authorization of COVID-19 messenger RNA (**mRNA**) vaccines in the United States (**US**) estimated the efficacy of a 2-dose series to be >90% in preventing symptomatic infection[1,2]. Post-authorization observational studies continued to estimate high vaccine effectiveness (**VE**) against symptomatic infection, even after the initial emergence of the SARS-CoV-2 Alpha and Delta variants[3]. However, by fall 2021 it was evident that waning immunity was contributing to decreased VE and increasing Delta variant incidence throughout the US. Subsequently, a booster dose was authorized and recommended for high-risk groups[4]. Recommendations for a booster dose were gradually expanded to include all people aged ≥12 years in the US as of December 30, 2022.

In contrast to previous SARS-CoV-2 variants of concern, the Omicron variant was demonstrated to have the capacity to evade immunity from prior infection shortly after it emerged in fall 2021[5]. Incidence of breakthrough infections among the vaccinated also increased following Omicron variant emergence, but VE against severe outcomes remained high, particularly among those who received a booster dose[6]. Less is known about the extent to which a booster dose protects against symptomatic Omicron infection.

During a period of Omicron circulation, we estimated relative VE of mRNA booster vaccination versus primary two-dose series in an ongoing community cohort in rural central Wisconsin, US.

## METHODS

The **P**rospective **A**ssessment of **C**OVID-19 in a **C**ommunity (**PACC**) study is an ongoing longitudinal cohort. PACC participants include persons <1 to >90 years of age who were randomly sampled and recruited from a defined community cohort in which nearly all residents receive care from the Marshfield Clinic Health System (**MCHS**). Participants completed a weekly symptom survey and were instructed to immediately report the onset of new respiratory symptoms. During the period of Omicron circulation, an anterior nasal swab was self- or parent-collected when participants reported ≥1 of the following: fever, cough, loss of smell or taste, sore throat, muscle/body aches, shortness of breath, or diarrhea. SARS-CoV-2 infection was defined as a positive real-time reverse transcription polymerase chain reaction result from a self-collected swab or a positive molecular clinical test result documented in the MCHS electronic health record. COVID-19 vaccination status was determined from MCHS electronic health records, the Wisconsin Immunization Registry, and review of participant vaccination cards. Participant characteristics including age, sex, and presence of chronic health conditions were determined by survey at enrollment.

This analysis included vaccinated PACC participants followed from 12/20/2021 through 02/24/2022 who were aged ≥12 years and were booster-dose eligible (≥5 months after completion of an mRNA vaccine primary series). During the follow-up period approximately 96% of SARS-CoV-2 viruses sequenced in Wisconsin were determined to be Omicron variant[7]. Hazard ratios comparing SARS-CoV-2 infection between booster versus primary series vaccinated individuals were estimated using age-adjusted Cox proportional hazards models with time-varying booster status. Primary series vaccinated person-time began 12/20/21 or ≥5 months after completion of the second dose, whichever was later, and ended at receipt of the third dose or end of study period. Booster dose vaccinated person-time began 12/20/21 or ≥14 days after receipt of the third dose, whichever was later. Relative VE of booster versus primary series vaccination was calculated as 100*(1 – adjusted hazard ratio). Sensitivity analyses estimated VE excluding participants 1) who reported that they were in an immunocompromised state, or 2) who had evidence of SARS-CoV-2 infection prior to 12/20/2021. Prior SARS-CoV-2 infection was defined as a positive molecular SARS-CoV-2 test result from research or clinical testing prior to 12/20/2021 or self-report at study enrollment.

This study was reviewed and approved by the Institutional Review Board at the Marshfield Clinic Research Institute. Participants, or parents of minor participants, provided informed consent prior to participation. Participating children ≥7 years also provided assent for participation.

## RESULTS

There were 884 participants included in the analysis. Participants ranged in age from 12 to >90 years (median: 57, IQR: 37 – 72), 62% were female, and 42% had ≥1 chronic health condition (Table 1). A total of 230 (26%) participants completed the primary series without boosting contributing 13,301 person-days to the analysis, and 654 (74%) received a booster dose by the end of follow-up contributing 37,550 person-days to the analysis. The median time from completion of the primary series to the start of follow-up (12/20/2021) was 233 days for unboosted participants and 275 days for boosted participants. The median time from booster dose to the start of follow-up was 33 days for boosted participants. Among unboosted individuals, 170 (74%) received 2 doses of BTN162b2 (Pfizer-BioNTech Comirnaty) vaccine, 60 (26%) received 2 doses of mRNA-1273 (Moderna Spikevax) vaccine, and none received mixed products as part of their primary series. Among boosted individuals, 406 (62%) received 2 doses of BTN162b2, 247 (38%) received 2 doses of mRNA-1273, and 1(0.2%) received mixed products as part of their primary series. Booster doses were 65% BTN162b2 and 35% mRNA-1273.

**Table 1.** Participant characteristics by SARS-CoV-2 vaccination and infection status: **P**rospective **A**ssessment of **C**OVID-19 in a **C**ommunity (**PACC**) study 12/20/2021 – 02/24/2022.

|  | Booster Vaccinated<sup>a</sup> |  |  | Primary Series Vaccinated ≥5 months<sup>b</sup> |  |  |
| --- | --- | --- | --- | --- | --- | --- |
|  | SARS-CoV-2 Infected<sup>c</sup><br>No. (%) | SARS-CoV-2 Uninfected<sup>c</sup><br>No. (%) | Total<br>No. (%) | SARS-CoV-2 Infected<sup>c</sup><br>No. (%) | SARS-CoV-2 Uninfected<sup>c</sup><br>No. (%) | Total<br>No. (%) |
| Age Group (years) |  |  |  |  |  |  |
| 12-17 | 0 (0) | 27 (4.4) | 27 (4.1) | 10 (21.7) | 35 (19.0) | 45 (19.6) |
| 18-49 | 11 (30.6) | 177 (28.6) | 188 (28.8) | 27 (58.7) | 75 (40.8) | 102 (44.4) |
| 50-64 | 11 (30.6) | 135 (21.8) | 146 (22.3) | 7 (15.2) | 49 (26.6) | 56 (24.4) |
| ≥65 | 14 (38.9) | 279 (45.2) | 293 (44.8) | 2 (4.4) | 25 (13.6) | 27 (11.7) |
| Sex |  |  |  |  |  |  |
| Female | 22 (61.1) | 383 (62.0) | 405 (61.9) | 31 (67.4) | 111 (60.3) | 142 (61.7) |
| Male | 14 (38.9) | 235 (38.0) | 249 (38.1) | 15 (32.6) | 73 (39.7) | 88 (38.3) |
| Chronic Condition(s)<sup>d</sup> |  |  |  |  |  |  |
| Yes | 16 (44.4) | 302 (48.9) | 318 (48.6) | 11 (23.9) | 43 (23.4) | 54 (23.5) |
| No | 20 (55.6) | 316 (51.1) | 336 (51.4) | 35 (76.1) | 141 (76.6) | 176 (76.5) |
| Immune Compromised<sup>e</sup> |  |  |  |  |  |  |
| Yes | 2 (5.6) | 25 (4.0) | 27 (4.1) | 1 (2.2) | 5 (2.7) | 6 (2.6) |
| No | 34 (94.4) | 593 (96.0) | 627 (95.9) | 45 (97.8) | 179 (97.3) | 224 (97.4) |
| Prior SARS-CoV-2 Infection<sup>f</sup> |  |  |  |  |  |  |
| Yes | 0 (0) | 120 (19.4) | 120 (18.4) | 1 (2.2) | 51 (27.7) | 52 (22.6) |
| No | 36 (100) | 498 (80.6) | 534 (81.7) | 45 (97.8) | 133 (72.3) | 178 (77.4) |

<sup>a</sup> Receipt of 3 doses of SARS-CoV-2 messenger RNA vaccine. Individuals are considered boosted ≥14 days after receipt of their third dose.
<sup>b</sup> Receipt of 2 doses of SARS-CoV-2 messenger RNA vaccine ≥5 months prior to 12/20/2021.
<sup>c</sup> SARS-CoV-2 infection status during the follow-up period 12/20/2021 – 02/24/2022.
<sup>d</sup> At enrollment, participants were asked to report any of the following chronic conditions: asthma, cancer, chronic kidney disease, COPD, hypertension, immunocompromised state, serious heart condition (heart failure, coronary artery disease or cardiomyopathies), or Type 2 diabetes.
<sup>e</sup> Immune compromised state self-reported by study participant at enrollment.

A total of 82 (9.3%) SARS-CoV-2 infections were identified, 46 occurred after receiving the primary series and 36 after a booster dose. This corresponded to SARS-CoV-2 incidence of 9.6/10,000 person-days among those receiving a booster dose and 34.6/10,000 person-days among those only receiving the primary series. Relative VE was estimated to be 66% (95% CI: 46%, 79%) favoring the booster dose compared to completion of only the primary vaccine series. SARS-CoV-2 infection prior to 12/20/2021 was identified in 120 (18%) boosted participants and 52 (23%) primary series participants. Reinfection during the analysis period was only identified in 1 primary series vaccinated participant out of the 172 total previously infected participants. Relative VE was 70% (95% CI: 51%, 81%) in sensitivity analyses excluding the 172 participants with prior infection. Relative VE was also similar when excluding participants in an immunocompromised state (67% [95% CI: 47%, 80%]).

## DISCUSSION

In our study, an mRNA vaccine booster dose added significant protection against symptomatic SARS-CoV-2 infection during a period of predominant Omicron circulation. As of March 2022, 70% of the US population ≥5 years of age are fully vaccinated[8]. Among those fully vaccinated individuals ≥12 years of age who are booster eligible, only 47% have received a booster dose. It is clear that COVID-19 vaccine booster doses have been very effective in preventing hospitalization and death, but uncertainty around effectiveness in preventing infection may contribute to vaccine hesitancy in some populations. Other than concerns of side-effects, concerns over vaccine effectiveness were the most commonly reported reasons for COVID-19 vaccine hesitancy in a large US cohort[9]. Our study provides important evidence that booster doses are effective in preventing Omicron infection in the community; these results can inform communications to encourage vaccination.

Given the timing of our study, most individuals who received a booster dose had done so recently. In contrast, the median time since completion of primary series was over 7 months for unboosted participants. Therefore, booster dose protection is likely achieved, in part, by boosting immunity that waned following primary series vaccination. However, it has also been demonstrated that serum from recently boosted individuals has greatly improved capacity to neutralize Omicron than serum from individuals who recently completed 2 dose primary series vaccination[10]. This suggests that an mRNA booster dose increases the breadth of response in addition to boosting the amount of circulating antibody. The exact mechanisms by which an mRNA vaccine booster dose might increase cross-neutralization relative to primary series are unclear.

A limitation of our study was that sample size was insufficient to estimate absolute VE relative to unvaccinated individuals. However, our relative VE estimate is similar to relative VE (66%) estimated by a large national pharmacy-based testing program that also found absolute VE of 67% against Omicron infection for 3 doses of mRNA vaccine[11]. Although absolute primary series VE was not estimated in that study, the nearly identical estimates of relative VE and absolute VE of a booster dose in that study suggest an absolute primary series VE near 1%. A similar study using community testing in England estimated an absolute mRNA booster VE between 64% and 74% against Omicron infection, depending on vaccine type and time since boost[12]. For 2-dose primary series alone, absolute VE was estimated 9% and 15% ≥25 weeks after BTN162b2 and mRNA-1273 vaccination, respectively. Taken together, the results of the current study and others suggest that the primary 2-dose mRNA vaccination provided little to no protection against Omicron infection in winter 2022 among those that did not receive a booster dose.

VE against symptomatic SARS-CoV-2 infection in the community provides additional support for current booster recommendations. Our results also further highlight the fact that vaccine-induced protection against SARS-CoV-2 infection is temporary and is impacted by waning immunity as well as evolution of circulating virus. As is the case with influenza, routine vaccination and regular evaluation of vaccine composition will be necessary to address ongoing SARS-CoV-2 transmission.

## Data Availability

All data produced in the present study are available upon reasonable request to the authors.

## ACKNOWLEDGEMENTS

We thank the following for their contributions to the study: Roxy Eibergen, Lynn Ivacic, Diane Kohnhorst, Erik Kronholm, DeeAnn Polacek, Carla Rottscheit, Elizabeth Armagost, Bobbi Bradley, Hannah Berger, Adam Bissonnette, Joshua Blake, Thomas G. Boyce, Keegan Brighton, Gina Burbey, Deanna Cole, Leila L. Deering, Cody DeHamer, Rachel Fernandez, Sherri Guzinski, Kayla Hanson, Erin Higdon, Jacob P. Johnston, Julie M. Karl, Taylor Kent, Burney A. Kieke Jr., Sarah Kohn, Sarah L. Kopitzke, Tamara Kronenwetter Koepel, Stacey Kyle, Eric LaRose, Kate Lassa, Carrie Marcis, Karen McGreevey, Nidhi Mehta, Daniel Miesbauer, Jennifer Moran, Pamela Mundt, Lisa Ott, Nan Pan, Cory Pike, Rebecca Pilsner, Martha Presson, Nicole Price, Mark Riley, Jacklyn L. E. Salzwedel, Juan Saucedo, Kristin Seyfert, Alex Slenczka, Elisha Stefanski, Robert Strenn, Sandra K. Strey, Melissa Strupp, and Krishna C. Upadhyay at Marshfield Clinic Research Institute; Yoshihiro Kawaoka, Gabrielle Neumann, David Pattinson, Lizheng Guan and Peter Jester at University of Wisconsin-Madison; Fatimah Dawood, Constance Ogokeh, Carrie Reed, and Douglas Slaughter at Centers for Disease Control and Prevention.

## DISCLAIMER

The findings and conclusions in this report are those of the author(s) and do not necessarily represent the official position of the Centers for Disease Control and Prevention (CDC).

